# Up front and open, shrouded in secrecy, or somewhere in between? A Meta Research Systematic Review of Open Science Practices in Sport Medicine Research

**DOI:** 10.1101/2023.03.30.23287959

**Authors:** Garrett S. Bullock, Patrick Ward, Franco M. Impellizzeri, Stefan Kluzek, Tom Hughes, Charles Hillman, Brian R. Waterman, Kerry Danelson, Kaitlin Henry, Emily Barr, Kelsey Healey, Anu M. Räisänen, Christina Gomez, Garrett Fernandez, Jakob Wolf, Kristen F. Nicholson, Tim Sell, Ryan Zerega, Paula Dhiman, Richard D. Riley, Gary S Collins

## Abstract

**Objective:** To investigate the extent and qualitatively synthesize open science practices within research published in the top five sports medicine journals from 01 May 2022 and 01 October 2022.

**Design:** Meta-research systematic review

**Data Sources:** MEDLINE

**Eligibility Criteria:** Studies were included if they were published in one of the identified top five sports medicine journals as ranked by Clarivate. Studies were excluded if they were systematic reviews, qualitative research, grey literature, or animal or cadaver models.

**Results:** 243 studies were included. The median number of open science practices met per study was 2, out of a maximum of 12 (Range: 0-8; IQR: 2). 234 studies (96%, 95% CI: 94-99) provided an author conflict of interest statement and 163 (67%, 95% CI: 62-73) reported funding. 21 studies (9%, 95% CI: 5-12) provided open access data. 54 studies (22%, 95% CI: 17-included a data availability statement and 3 (1%, 95% CI: 0-3) made code available. 76 studies (32%, 95% CI: 25-37) had transparent materials and 30 (12%, 95% CI: 8-16) included a reporting guideline. 28 studies (12%, 95% CI: 8-16) were pre-registered. 6 studies (3%, 95% CI: 1-4) published a protocol. 4 studies (2%, 95% CI: 0-3) reported the availability of an analysis plan. 7 studies (3%, 95% CI: 1-5) reported patient and public involvement.

**Conclusion:** Sports medicine open science practices are extremely limited. The least followed practices were sharing code, data, and analysis plans. Without implementing open practices, barriers concerning the ability to aggregate findings and create cumulative science will continue to exist.

**What is already known:**

- Open science practices provide a mechanism for evaluating and improving the quality and reproducibility of research in a transparent manner, thereby enhancing the benefits to patient outcomes and society at large.
- Understanding the current open science practices in sport medicine research can assist in identifying where and how sports medicine leadership can raise awareness, and develop strategies for improvement.

**What are the new findings:**

- No study published in the top five sports medicine journals met all open science practices
- Studies often only met a small number of open science practices
- Open science practices that were least met included providing open access code, data sharing, and the availability of an analysis plan.

## Introduction

Advances in how we plan and conduct research has created (in principle) opportunities for international collaboration^1^ and sharing of data that lead to gains in overall scientific knowledge,^2-4^ that was unimaginable only a few decades ago. While sports medicine and science have paralleled these scientific attainments,^5-7^ there is still much to learn in terms of preventing detrimental sport medical outcomes and improving athlete health.^5^

It is highly likely that some progress in sport medicine and science is being stymied as a result of significant and serious barriers that have been recently highlighted in the current literature. For example, numerous systematic literature reviews have consistently demonstrated the use of suboptimal methods and incomplete reporting,^8^ indicating a risk of adopting findings based on misleading or potentially harmful conclusions.^8 9^ Methodological flaws and misconduct such as ‘p-hacking’ and hypothesizing after the results are known (HARKing)^10^ further impede scientific advancements by inflating risk of type 1 error.^10 11^ Incomplete reporting and a lack of transparency in the scientific design, conduct and reporting of scientific studies, including unavailability of protocols, analysis plans, code, and data enables these practices to continue.

This may also, in combination with poor, or incomplete reporting of final studies, restrict the ability to identify valid findings from well-designed studies or limit the accuracy of aggregated analyses.^12 13^ In addition, research conduct is often limited in sport because of sample size restrictions that are exerted by using datasets from individual teams or organizations.^14^ While data sharing initiatives can overcome this barrier, a team’s proprietary data is often strongly protected.^15^

One overriding principle to help improve transparency and utility in research is through *open science*.^16^ Open science is a movement to make all scientific materials and results accessible to all levels of society^17^ and is a process where scientists openly share protocols and analysis plans, register studies, report results, and share data and code.^18^ To advance science, and within sports medicine, to continuously improve athlete health, the entire scientific process must be based on the Open Science paradigm and must be “findable, accessible, interoperable, and reusable (FAIR)”.^19^ This process allows fellow scientists to evaluate, replicate, and confirm previous research from transparent methods, open data and code.^16 19^

While open science practices have been relatively well adopted in the physical and biological sciences,^5^ however, these fields do not have the same ethical considerations as the medical sciences.^2 5 18^ Sports medicine and science has further concerns such as competition between clubs and athlete re-identification which may constitute a barrier to open science uptake.^16^ Despite these issues, funders and charity organizations increasingly require plans for open science practices to be embedded in grant applications for funded sports medicine research.^2 5 18^

Adopting open science practices can improve research transparency and credibility and advance athlete care and health at a faster pace compared to opaque (i.e., less transparent) research.^16^ However, it is not clear as to what extent open science practices are adopted within sports medicine and science research. Understanding the current state can assist academics, practitioners, journal editors, reviewers, and funding bodies in determining where and how they can employ and improve open science practices, potentially accelerating collaboration, methodological transparency, and athlete health outcomes.^16 20 21^ Therefore, the purpose of this study was to carry out meta-research using a systematic review to investigate the extent and qualitatively synthesize open science practices within research published in the top five sports medicine journals from 01 May 2022 to 01 October 2022.

## Methods

### Study design

The design of this meta research systematic review was informed by previous work by Hardwicke et al.^22^ This study was reported using guidelines for reporting methodology research^23^ and the Preferred Reporting Items for Systematic Reviews and Meta-Analyses Protocol (PRISMA-P).^24^ Evaluation of open science practice was informed through two sources, Evaluating implementation of the Transparency and Openness Promotion (TOP) guidelines^25^ and the review by Tennant et al.^26^ This study was registered on the Open Science Framework (https://osf.io/4amek/). The final draft manuscript was uploaded and made available on a pre-print server prior to peer review.

### Relevant party involvement (i.e., Patient and public involvement)

The research question was developed following consultation with several professional groups, including non-academic partners and individuals who had an interest in or were affected by amateur, collegiate and professional sport. These groups included physiotherapists, physicians, sports performance coaches, athletic trainers, as well as statistical and methodological researchers. These groups also met virtually to discuss strategy and study progress, preliminary results and interpretation of findings, and provide input into the plan for dissemination of findings

### Equity, Diversity, and Inclusion

After consideration of the necessity to involve relevant parties and collaborators with required expertise, the author team consists of a diverse range of individuals, including students, clinicians, and early, middle, and late career researchers with balance from people whom identify as men and women, different age groups, and nationalities.

### Study eligibility criteria

Article inclusion and exclusion criteria are reported in Table 1.

**Table 1.** Article Inclusion and Exclusion Criteria

| **Inclusion Criteria** | **Exclusion Criteria** |
| --- | --- |
| Studies published in one of the identified top five sports medicine journals ranked by Clarivate journal citation rankings: (1) *British Journal of Sports Medicine*, (2) *Journal of Sport and Health Science*, (3) *American Journal of Sports Medicine*, (4) *Medicine Science Sport and Exercise*, (5) *Sports Medicine-Open*) | Systematic reviews, scoping reviews, meta-analysis |
| Studies published in special edition journal issues | Qualitative Research |
| Original scientific research published as a full peer reviewed paper | Case reports, editorials, letters to the editor |
| Randomized control trials, observational studies | Grey literature |
| Published in English | Studies using animal and cadaver models |

### Search strategy and journal selection

Sports medicine journals were chosen based on Clarivate journal citation rankings. While these rankings have limitations,^27 28^ this method was chosen to remove author subjectivity, and therefore prohibited the author team from selectively cherry picking journals. After excluding journals that are focused on systematic reviews (*Sports Medicine*; *Exercise Immunology Review*) and qualitative research (*Qualitative Research in Sport, Exercise and Health*), the top five journals were (1) *British Journal of Sports Medicine*, (2) *Journal of Sport and Health Science*, (3) *American Journal of Sports Medicine*, (4) *Medicine Science Sport and Exercise*, and (5) *Sports Medicine-Open*. These five journals were searched through MEDLINE on October 10, 2022 for all articles published over a six-month time period, between May 1, 2022 and October 1, 2022 (Appendix 1).

### Study Selection

All reviewers participated in an online training session (led by GB) that provided information for article screening and the data extraction process. A calibration exercise was then performed prior to screening, with all reviewers required to achieve greater than 90% agreement prior to screening. Titles and abstracts were screened independently for eligibility in equal numbers of randomized articles by paired screening groups (PW and FI, TH and CH, KD and KH, EB and KH, AR and CG, GF and JW, TS and RZ). The full-text of eligible studies were then recovered and screened independently by the same screening pairs.^29^ Title and abstract and full-text study disputes were resolved by consensus within each screening pair. If consensus could not be resolved, the lead author (GB) had final resolution on study inclusion or exclusion. Selected full-text articles were retrieved through university online library portals. If a study could not be retrieved, the authors were contacted to request full text, and, if required, interlibrary loan with the assistance of a librarian was attempted. If a full-text article could not be retrieved, the study was excluded from the review.^29-31^ All screening was performed in Covidence systematic review software (Veritas Health Innovation, Melbourne, Australia).

### Data Extraction

Data were extracted by the same screening pairs (PW and FI, TH and CH, KD and KH, EB and KH, AR and CG, GF and JW, TS and RZ), entered into a customized electronic database, using the recommended practices of The National Institute for Health and Care Excellence evidence tables.^32 33^ Each pair independently extracted data, with conflicts resolved first by consensus, followed by the lead author (GB). A random sample of three articles from each data extraction team were screened and graded by the study leads (GB, GC) for quality control. Data extraction included author details (e.g., first author surname, title, study design, journal, month of publication, and sport. Open science methods were extracted in accordance with the TOP guidelines,^25^ with further open science data comprising patient public involvement (Table 2).^26^ Any articles that were electronic publications ahead of print were extracted, but were not complicit to open science criteria such as disclosing author conflicts or reporting funding. The Open Science data were extracted as a ‘yes’ or ‘no’ for meeting the criteria.

**Table 2.** Open Science Practices evaluated in the review (*adapted from the TOP guidelines^25^)

| **Open Science Practice** | **Criterion** |
| --- | --- |
| 1. Conflict of Interest Statement | Manuscript provides details on any author conflicts of interest. |
| 2. Funding Statement | Manuscripts describe funding, and the role of any funders. |
| 3. Data Citation | Manuscript provides details on the provenance of data, with a clear identifier (e.g., digital object identifiers, website, or link to digital repository). |
| 4. Data Transparency | Manuscript states where any data are available (e.g., in a data sharing statement), such as a data warehouse or repository, and where to access them through an embedded link. May be within manuscript, or as a separate section (i.e., data availability statement). |
| 5. Analysis Code Transparency | Manuscript includes details on code availability (i.e., in supplementary materials, or has an available link to a repository within the manuscript). |
| 6. Materials Transparency | Manuscript state where any materials (such as patient reported outcomes or survey questions) are available, e.g., included as an appendix or a link to a repository. |
| 7. Design & Analysis Reporting Guideline | Manuscript cites and claims use of an appropriate reporting guideline. |
| 8. Study Registration | Manuscripts state study registration number with an open access database (e.g., Prospero, clinictrials.gov). |
| 9. Study Protocol | Manuscripts states a study protocol was available in an open access repository (e.g., Open Science Framework) or published in an open access journal. |
| 10. Statistical Analysis Plan | Manuscripts states a statistical analysis plan was available in an open access repository (e.g., Open Science Framework) or published in an open access journal. |
| 11. Patient & Public Involvement | Manuscript describes any patient and public involvement, also known as ‘citizen science’. |
| 12. Replication | Replication studies, such as registered reports are performed to validate previous research. |

## Data Sharing

The reconciled extracted data that form the results in this study are available in the Open Science Framework (https://osf.io/4amek/).

### Collating, Summarizing and Reporting the Results

Overall screening agreement and quality control agreement were calculated by Cohen’s Weighted Kappa. Proportions, point estimates and 95% confidence intervals were summarized across all included studies, calculated according to whether or not each criterion for open science practice was met in each study. Data were also stratified according to journal, study design and sport. Data were further summarized through median, range, and interquartile range (IQR) of articles meeting open science practices Due to small sample size and proportions at or around zero, Clopper-Pearson confidence intervals were calculated for proportions.^34^ A narrative synthesis was performed. All analyses were performed in R 4.02 (R Core Team (2021). R: A language and environment for statistical computing. R Foundation for Statistical Computing, Vienna, Austria. URL https://www.R-project.org/). The *dplyr* package was used for cleaning and calculations.

## Code Sharing

Analytical code used to summarize the findings in this paper are available on the Open Science Framework (https://osf.io/4amek/).

## Results

A total of 361 titles and abstracts were identified over the 6-month sample period for the five sports medicine journals. After removal of duplicates, the screening process identified 243 studies that met inclusion criteria and so were included in the review (Figure 1). Overall, the Kappa agreement between reviewers for data extraction was 0.86. Random sample quality control agreement was 0.98.

**Figure 1.**
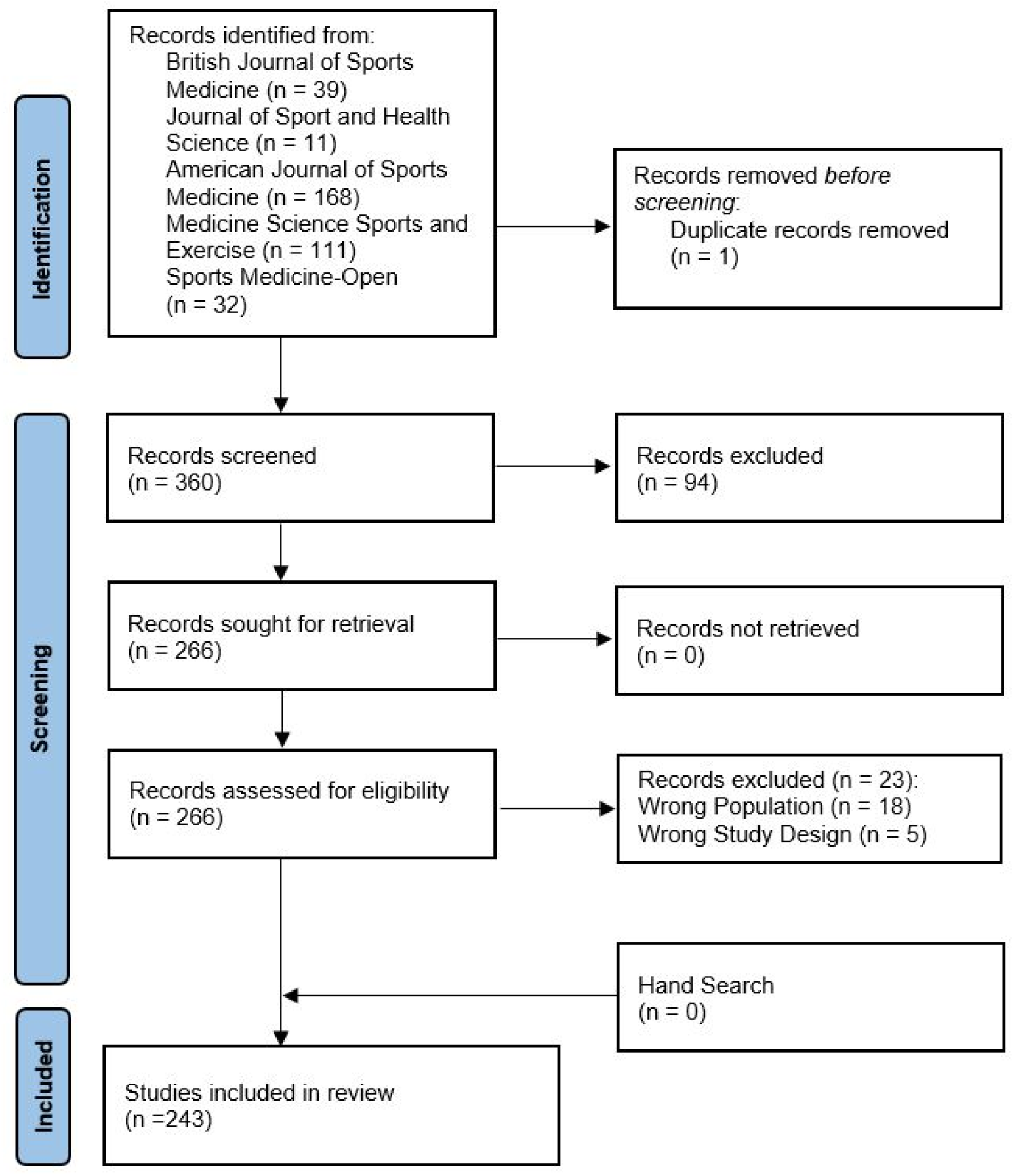
PRISMA Flow Diagram

### Study Characteristics

Of the 243 included studies, a total of 20 (8%, 95% CI: 5-12) were included from the British Journal of Sports Medicine, 5 (2%, 95% CI: 0-7) from Journal of Sport and Health Science, 112 (46%, 95% CI: 40-53) from the American Journal of Sports Medicine, 85 (35%, 95% CI: 29-41) from Medicine Science Sport and Exercise, and 21 (9%, 95% CI: 5-13) from Sports Medicine-Open.

A total of 94 studies (39%, 95% CI: 33-45) were prospective cohort studies, 58 (24%, 95% CI: 19-30) retrospective cohort, 32 (13%, 95% CI: 9-18) cross-sectional, 29 (12%, 95% CI: 8-17) were randomized controlled trials, 14 (6%, 95% CI: 3-9) case-control, 14 (6%, 95% CI: 3-9) case series, 1 (<1%, 95% CI: 0-2) quasi-experimental, and 1 (<1%, 95% CI: 0-2) economic and decision analysis.

A total of 81 studies (33%, 95% CI: 27-40) investigated general population exercise, 57 (23%, 95% CI: 18-29) multiple sports, 51 (21%, 95% CI: 16-27) general orthopaedic patients, 15 (6%, 95% CI: 3-10) running, 10 (4%, 95% CI: 2-7) baseball, 4 (2%, 95% CI: 1-4) cycling, 4 (2%, 95% CI: 1-4) military, 3 (1%, 95% CI: 0-4) soccer, 3 (1%, 95% CI: 0-4) swimming and diving, 2 (1%, 95% CI: 0-3) football, and 1 (<1%, 95% CI: 0-2) for individual sports of basketball, e-sports, handball, lacrosse, motor sports, netball, occupational population, pregnant athletes, rowers, and skiing.

### Evaluation of Overall Open Science Practices

No studies met all open science practices. One study (<0.1%, 95% CI: 0-2) met at least 8 out of 12 open science criteria. The median number of open science practices met per study was 2 (range: 0-8; IQR: 2). Please refer to supplementary data (https://osf.io/4amek/) for individual study evaluations.

A total of 234 (96%, 95% CI: 93-98) reported author conflicts, and 163 (67%, 95% CI: 61-73) provided details on funding. A total of 21 (9%, 95% CI: 5-13) provided open access data through an embedded a link or made data available in the supplementary material. Fifty-four (22%, 95% CI: 17-28) included a data availability statement or signposted where data was available. Of these 54 studies, 39 (72 %, 95% CI: 58-84) reported data was available upon reasonable request, and 15 (28%, 95% CI: 16-42) reported a publicly available site to request data. Three studies (6%, 95% CI: 1-15) provided a link, made available the supplementary material, or highlighted where open access code was available.

Seventy-six studies (32%, 95% CI: 22-34) had fully transparent and available materials and methods. Twenty-eight studies (12%, 95% CI: 8-16) reported following a reporting guideline. Of these, 14 (50%, 95% CI: 31-69) reported the Consolidated Standards of Reporting Trials (CONSORT) guidelines,^35^ 11 (39%, 95% CI: 22-59) reported the Strengthening the Reporting of Observational Studies in Epidemiology (STROBE) guidelines,^36^ 4 (14%, 95% CI: 4-33) the TRIPOD guidelines,^37^ and 1 (4%, 95% CI: 0-18) the Checklist for Reporting Results of Internet E-Surveys (CHERRIES) guidelines.^38^ Twenty eight studies (12%, 95% CI: 8-16) reported preregistration and 6 (3%, 95% CI: 1-5) published a protocol in an open access journal or placed it in an open science repository. Four (2%, 95% CI: 0-4) reported the availability of an analysis plan. No studies (0%, 95% CI: 0-2) were replication studies. Seven studies (3%, 95% CI: 1-6) reported patient and public involvement or citizen science. (Figure 2).

**Figure 2.**
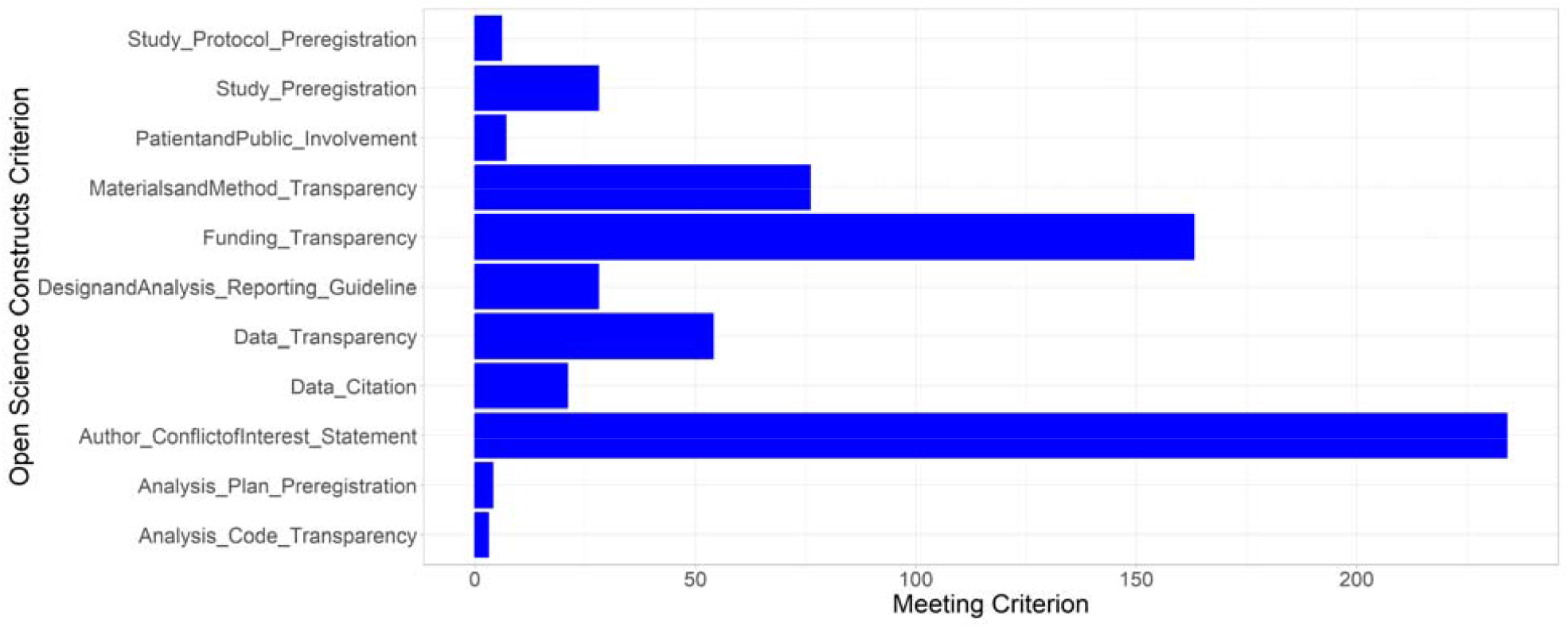
Breakdown of Open Science Practice

**Figure 3.**
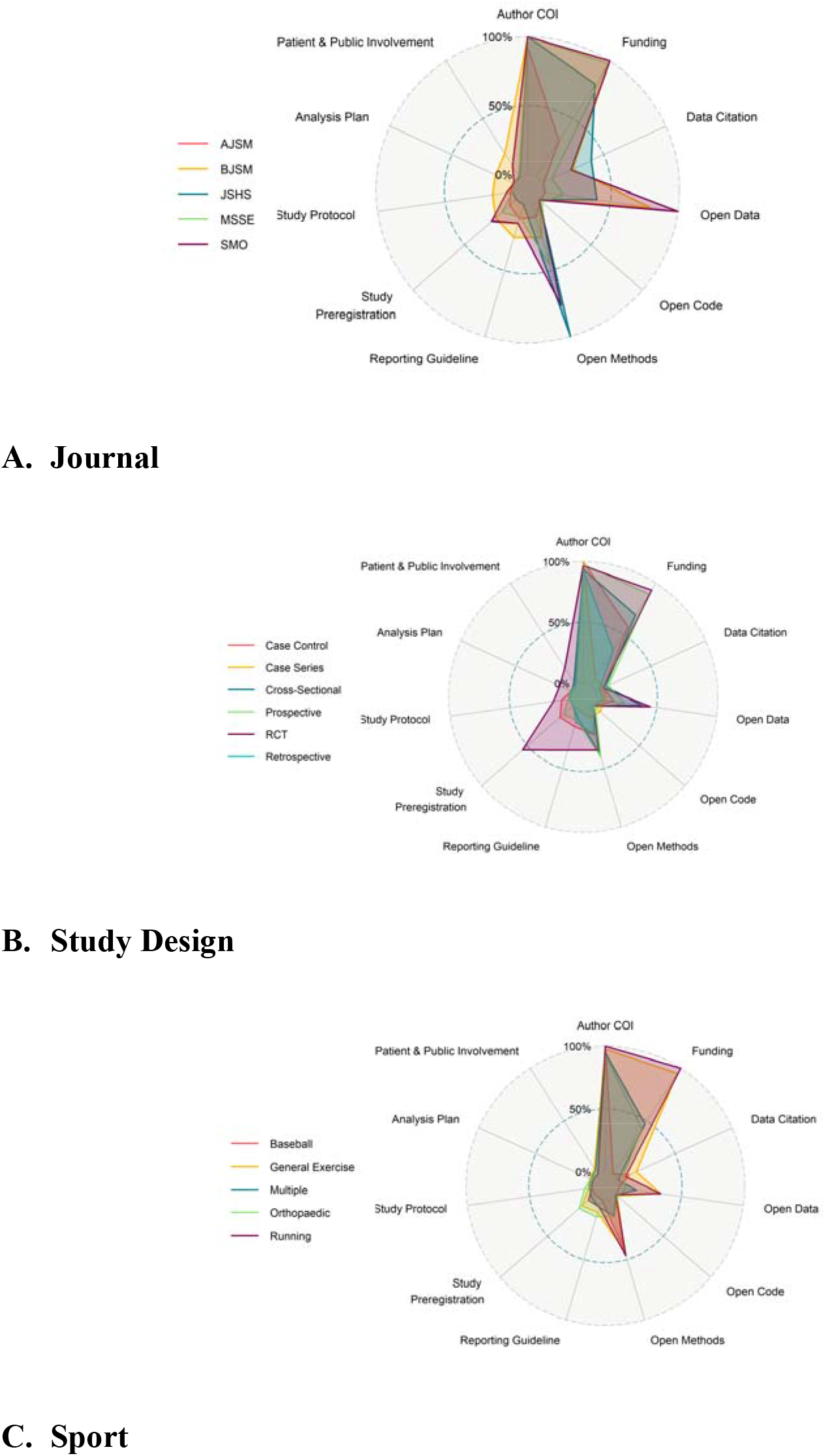
Open Sciences Practices *Replication is not reported as no studies were replication COI = Conflict of Interest. AJSM = American Journal of Sports Medicine, BJSM = British Journal of Sports Medicine, JSHS = Journal of Sport and Health Science, MSSE = Medicine and Science in Sport and Exercise, SMO = Sports Medicine-Open. *Open Science Practices by Journal*

The median number of open science practices met per article for British Journal of Sports Medicine was 3 (range: 2-8; IQR: 3), the median for Journal of Sport and Health Science was 3 (range: 3-5; IQR: 1), the median for American Journal of Sports Medicine was 1 (range: 1-7; IQR: 1), the median for Medicine Science Sport and Exercise was 2 (range: 0-6; IQR: 1), and the median for Sports Medicine-Open was 4 (range: 3-7; IQR: 1).

Greater than 50% of studies published in each journal reported author conflicts and funding. Less than 40% of studies reported for data citation in each journal, and only two journals (AJSM and MSSE) had any articles report open access code. The use of reporting guidelines was reported in 25% or less of studies published in each journal. Only studies in two journals (AJSM and BJSM) reported the availability of statistical analyses plans. Studies in the British Journal of Sports Medicine were twice as likely to report patient and public involvement (Figure 2A; Appendix 2)

### Open Science Practices by Study Design

The median number of open science practices met per study for prospective cohorts was 2 (range: 0-6; IQR: 2), the median for randomized controlled trials was 4 (range: 1-8; IQR: 2), the median for retrospective cohorts was 1 (range: 1-4; IQR: 1), the median for case-controls was 2 (range: 1-7; IQR: 2), the median for cross-sectional studies was 2 (range: 1-5; IQR: 3), the median for case series was 1 (range: 1-6; IQR: 0). Economic and decision analyses and quasi-experimental studies both only included one study.

All study designs had similar percentage in terms of meeting the open science criteria for author conflicts, funding, data transparency, and analysis and code transparency. Randomized controlled trials had four times greater percentage of studies that used reporting guidelines and five times greater percentage for registering a study. Randomized controlled trials had three times greater percentage for reporting availability of a statistical analysis plan, and five times greater percentage for reporting patient and public involvement (Figure 2B; Appendix 2)

### Open Science Practices by Sport

The median number of open science practices met per study for general population exercise was 2 (range: 0-8; IQR: 2), the median for multiple sports was 1 (range: 1-5; IQR: 1), the median for general orthopaedic patients was 2 (range: 1-7; IQR: 1), the median for running was 3 (range: 1-5; IQR: 2), and the median for baseball was 1 (range: 1-3; IQR: 1).

All sport, exercise, and orthopaedic population studies demonstrated a similar percentage for meeting open science criterion for author conflicts, funding, data transparency, analysis and code transparency, study registration, analysis plan, and patient and public involvement. Studies that investigated orthopaedic patients had two times greater percentage of using a reporting guideline compared to studies that studied investigated sport and exercise populations (Figure 2C; Appendix 2).

## Discussion

The primary finding of this study was that no studies from the top five sports medicine journals met all open science practices. One study met 8 out of 11 open science practices, whereas the median number of open science practices met was only two. Open science practices that were least likely to be upheld were sharing of analysis code, sharing data, and availability of an analysis plan. When stratifying by study design, randomized controlled trials reported adopting the most open science practices criteria, whilst observational studies the least.

Our review revealed that the severely limited use of open science practices which has significant implications for the analysis and impact of research findings on clinical and sports science practice. Current sport medicine literature has generally demonstrated a high risk for poor research practices and publication bias towards selecting ‘statistically significant’ findings,^39 40^ with over 80% of sports medicine research resulting in confirmed hypotheses.^40^ This is an inordinate high positive rate, but when considering studies that are of high methodological and reporting quality and of adequately powered studies, this resulted in less than 50% ‘significant’ findings.^41 42^

This indicates substantial potential for bias, both publication bias as well as potential shortcomings in the design, execution, or analysis resulting in the inability to replicate studies also impedes the generalizability of results.^43^ In addition, the limited adoption of open science practices makes it challenging to test reproducibility and generalizability of the published results. The importance of study replication has been highlighted previously, whereby an open science initiative replicated 100 psychological studies that reported ‘statistically significant’ results, with only 37% reporting positive results after replication.^44^ The implausibly high prevalence of statistically significant results is detrimental to evidence-based practice.^43^ False positives(a ‘statistically significant’ result, when in reality no effect exists) could be used to justify the identification of risk factors or use of potential interventions that clinicians and organizations invest time and resources to implementing, with no effect or possibly a harmful effect. Without improved and consistent open science uptake and research integrity, sports medicine research will continue to be limited.

Of particular importance, our review found that sports medicine and science studies demonstrated a paucity in sharing open access analysis code, data, and availability of analytical plans. Freely accessible statistical code and data sharing offers opportunities to other researchers to replicate statistical methods and results,^45 46^ it can also facilitate the reporting of errors,^16 21^ aggregate findings,^47 48^ and combine data from different sources to answer research questions that can’t be answered using single datasets.^16 21^ Unavailability of code and data hinders the sports medicine community’s ability to confirm results and combine data, to improve cumulative science. ^47^ It should be noted that while a number of studies reported their data is available upon request statement, which was technically meeting specific open science criteria, this statement is woefully inadequate, and has not resulted in increased access to data within the greater scientific literature.^49^ Thus, the overall prevalence of open data is likely lower than the reported results.

Randomized controlled trials demonstrated better adoption of open science practices compared to other study designs. Randomized controlled trials are required to register protocols before study recruitment prior at registries such as clinicaltrials.gov. Further, many journals, require RCTs to submit Consolidated Standards of Reporting Trials (CONSORT)^35^ checklists at the time of manuscript submission. The stricter study registration and methodological reporting of RCTs is due to the inherent risk, and thus patient protection required. Other methodological designs used in sport medicine, most notably observational studies, should require the same registration and methodological rigor, as these studies also inform evidence-based practice.^50^

As demonstrated within this meta-research systematic review, the reported open science practices are limited for studies published across the top five ranked journals within sports on medicine and science in the Clarivate journal citation rankings. While this is only a 6-month sample of selected sports medicine and science journals, it is possible that open science practice in other sports medicine journals may be even more limited, due to the smaller scientific barriers attributed to lower ranking journals.^51^

Based on the findings of this review, we strongly argue that the sports medicine community and journal editorial boards should make open science practices a priority before publication. Mandating study registration, availability of protocols, analytical plans, data, open access code, and requiring reporting author conflicts of interest, funding, and guideline checklists at submission are low hanging fruit, which can be easily implemented across all journals. The practices should also be viewed as just doing good science.^52 53^ Reporting patient public involvement, also known as citizen science, is an easy accessible open science practice that can and should be mandated across all journals. While there may be special concerns about sharing sports medicine data,^16 21^ many of these barriers can be circumvented, as already shown through other biomedical scientific fields.^2 5 18^ Potential solutions include creating synthetic (i.e., simulated) data that mirrors the characteristics of the actual data, creating a gatekeeper warehouse for data access, and using federated access (i.e., data is housed and analyzed only within the data owner’s servers). Nevertheless, there is no current consensus on the barriers and facilitators or legal ramifications of open access data within sport, and there is a need and opportunity to engage all relevant parties in this discussion.

There were limitations to this study. This study only included the top five sports medicine journals, as ranked by Clarivate. Other sports medicine journals may demonstrate different open science practices. A 6-month sample was taken from these journals, which may decrease the overall precision in these findings, however this was a contemporary refection of current open science practice. Scoping reviews are broad in nature, which decreases the precision of specific scientific questions.

## Conclusions

Less than 20% of recommended open science practices were currently met by studies published in the top five sports medicine journals. Sharing code, data, and availability of analysis plans were the least followed open science practices. Randomized controlled trials revealed better adherence to open science practices compared to observational studies. The sports medicine research community, including journals, researchers, and relevant parties (i.e., patient public involvement) should prioritize open science practices before publication. Without implementing these practices, trust in methods and results will be compromised, thereby negatively impacting how the literature influences evidence-based practice. Understanding the barriers, particularly those associated legality of data-sharing, is likely essential to advancing this progress.

## Supporting information

PRISMA-ScR Checklist

## Data Availability

Data is available at https://osf.io/4amek/
Code is available at https://osf.io/4amek/

## Contributions

GSB and GSC conceived the study idea. GB and GC designed the study. GB and GC wrote the initial draft. GB, PW, FI, SF, TH, CH, BW, KD, KH, EB, KH, AR, CG, GF, JW, KN, TS, RZ, PD, RR, GC critically revised the manuscript. GB, PW, FI, SF, TH, CH, BW, KD, KH, EB, KH, AR, CG, GF, JW, KN, TS, RZ, PD, RR, GC approved the manuscript.

## Data Availability

Data is available at https://osf.io/4amek/

## Code Availability

Code is available at https://osf.io/4amek/

