## Supplement for "Up front and open, shrouded in secrecy, or somewhere in between? A Meta Research Systematic Review of Open Science Practices in Sport Medicine Research"

**Appendix 1.** Search Strategy

***British Journal of Sports Medicine****, 10/10/2022---39 results:*

*(((("British Journal of Sports Medicine"[Journal]) AND (("2022/05/01"[Date - Entry]: "2022/10/06"[Date - Entry]))) NOT (Systematic Review)) NOT (Editorial)) NOT (Letter to the Editor)* NOT (Narrative) NOT (Meta-Analysis)

***Journal of Sport and Health Science****, 10/10/2022---11 results:*

*(((("Journal of Sport and Health Science"[Journal]) AND (("2022/05/01"[Date - Entry]: "2022/10/06"[Date - Entry]))) NOT (Systematic Review)) NOT (Editorial)) NOT (Letter to the Editor)* NOT (Narrative) NOT (Meta-Analysis)

***The American Journal of Sports Medicine****, 10/10/2022---168 results:*

(((("The American journal of sports medicine"[Journal]) AND (("2022/05/01"[Date - Entry]: "2022/10/06"[Date - Entry]))) NOT (Systematic Review)) NOT (Editorial)) NOT (Letter to the Editor) NOT (Narrative) NOT (Meta-Analysis)

***Medicine and Science in Sport and Exercise****, 10/10/2022---111 results:*

(((("Medicine and science in sports and exercise"[Journal]) AND (("2022/05/01"[Date - Entry]: "2022/10/06"[Date - Entry]))) NOT (Systematic Review)) NOT (Editorial)) NOT (Letter to the Editor) NOT (Narrative) NOT (Meta-Analysis)

***Sports Medicine-Open****, 10/10/2022---32 results:*

(((("Sports Medicine-Open"[Journal]) AND (("2022/05/01"[Date - Entry]: "2022/10/06"[Date - Entry]))) NOT (Systematic Review)) NOT (Editorial)) NOT (Letter to the Editor) NOT (Narrative) NOT (Meta-Analysis)

**Appendix 2.** Open Science Criterion Stratified by Journal, Study Design, and Sport and Exercise

|  | | Conflict of Interest | Funding | Data Citation | Data Transparency | Analysis & Code Transparency | Materials & Methods Transparency | Reporting Guideline | Study Registration | Study Protocol | Analysis Plan | Patient & Public Involvement |
| --- | --- | --- | --- | --- | --- | --- | --- | --- | --- | --- | --- | --- |
|  | Stratified by Journal | | | | | | | | | | | |
| BJSM  (n = 20) | | 20  (100) | 20  (100) | 5  (25, 9-49) | 16  (80, 56-94) | 0  (0, 0-10) | 5  (25, 9-49) | 5  (25, 9-49) | 4  (20, 6-44) | 3  (15, 3-38) | 3  (15, 3-38) | 4  (20, 6-44) |
| JSHS (n = 5) | | 5  (100) | 4  (80, 28-99) | 2  (40, 0-83) | 2  (40, 0-83) | 0  (0, 0-36) | 5  (100) | 0  (0, 0-36) | 0  (0, 0-36) | 0  (0, 0-36) | 0  (0, 0-36) | 0  (0, 0-36) |
| AJSM  (n = 112) | | 103  (92, 85-96) | 35  (31, 23-41) | 2  (2, 0-6) | 2  (2, 0-6) | 1  (1, 0-5) | 11  (10, 5-17) | 12  (11, 6-18) | 7  (6, 3-12) | 2  (2, 0-6) | 1  (1, 0-5) | 0  (1, 0-3) |
| MSSE  (n = 85) | | 85  (100) | 83  (98, 92-100) | 7  (8, 3-16) | 13  (15, 8-25) | 2  (2, 0-8) | 39  (46, 35-57) | 8  (9, 4-18) | 12  (14, 8-23) | 0  (0, 0-3) | 0  (0, 0-3) | 1  (1, 0-6) |
| Sport Med-Open (n = 21) | | 21  (100) | 21  (100) | 5  (24, 8-47) | 21  (100) | 0  (0, 0-10) | 16  (76, 53-92) | 3  (14, 3-36) | 5  (24, 8-47) | 1  (5 , 0-24) | 0  (0, 0-10) | 2  (9, 1-30) |
|  | Stratified by Study Design | | | | | | | | | | | |
| RCT (n = 29) | | 28  (97, 92-100) | 27  (93, 77-99) | 2  (7, 1-23) | 13  (45, 26-64) | 0  (0, 0-8) | 10  (34, 18-54) | 11  (38, 21-58) | 16  (55, 36-74) | 4  (14 , 4-32) | 3  (10, 2-27) | 5  (17, 6-36) |
| Prospective Cohort  (n = 94) | | 92  (98, 93-100) | 84  (89, 81-95) | 11  (12, 6-20) | 22  (23, 15-33) | 2  (2, 0-7) | 38  (40, 30-51) | 8  (9, 4-16) | 10  (11, 5-19) | 1  (1, 0-6) | 1  (1, 0-6) | 1  (1, 0-6) |
| Retrospective Cohort  (n = 58) | | 54  (93, 83-98) | 20  (34, 22-48) | 2  (3, 0-12) | 3  (5, 1-14) | 0  (0, 0-4) | 12  (21, 11-33) | 5  (9, 3-19) | 0  (0, 0-4) | 0  (0, 0-4) | 0  (0, 0-4) | 0  (0, 0-4) |
| Cross-Sectional  (n = 32) | | 30  (94, 79-99) | 22  (69, 50-84) | 3  (9, 2-25) | 12  (38, 21-56) | 0  (0, 0-7) | 1  (3, 1-21) | 2  (6, 0-15) | 0  (0, 0-7) | 0  (0, 0-7) | 0  (0, 0-7) | 1  (3, 0-16) |
| Case-Control  (n = 14) | | 14  (100) | 8  (57, 29-82) | 1  (7, 0-34) | 2  (14, 2-43) | 0  (0, 0-15) | 3  (21, 5-51) | 2  (14, 2-43) | 2  (14, 2-43) | 1  (7, 0-34) | 0  (0, 0-15) | 0  (0, 0-15) |
| Case Series  (n = 14) | | 14  (100) | 1  (7, 0-34) | 1  (7, 0-34) | 1  (7, 0-34) | 1  (7, 0-34) | 1  (7, 0-34) | 0  (0, 0-15) | 0  (0, 0-15) | 0  (0, 0-15) | 0  (0, 0-15) | 0  (0, 0-15) |
| Quasi-Experimental  (n = 1) | | 1  (100) | 1  (100) | 1  (100) | 1  (100) | 0  (0, 0-90) | 0  (0, 0-90) | 0  (0, 0-90) | 0  (0, 0-90) | 0  (0, 0-90) | 0  (0, 0-90) | 0  (0, 0-90) |
| Economic & Decision Analyses  (n = 1) | | 1  (100) | 0  (0, 0-90) | 0  (0, 0-90) | 0  (0, 0-90) | 0  (0, 0-90) | 0  (0, 0-90) | 0  (0, 0-90) | 0  (0, 0-90) | 0  (0, 0-90) | 0  (0, 0-90) | 0  (0, 0-90) |
|  | Stratified by Sport & Exercise | | | | | | | | | | | |
| General Population Exercise  (n = 81) | | 79  (98, 91-100) | 77  (95, 88-99) | 13  (16, 9-26) | 27  (33, 23-45) | 1  (1, 0-7) | 35  (43, 32-55) | 9  (11, 5-20) | 11  (14, 7-23) | 2  (2, 0-9) | 2  (2, 0-9) | 4  (5, 1-12) |
| Multiple Sports  (n = 57) | | 54  (95, 85-99) | 27  (47, 34-61) | 0  (0, 0-4) | 8  (14, 6-26) | 0  (0, 0-4) | 8  (14, 6-26) | 4  (7, 2-17) | 2  (4, 0-12) | 1  (2, 0-9) | 1  (2, 0-9) | 2  (4, 0-12) |
| General Orthopaedic Patients  (n = 51) | | 50  (98, 89-100) | 23  (45, 31-60) | 2  (4, 1-13) | 3  (6, 1-16) | 1  (2, 0-10) | 8  (16, 7-29) | 8  (16, 7-29) | 9  (18, 24-52) | 3  (6, 1-16) | 1  (2, 0-10) | 0  (0, 0-4) |
| Running  (n = 15) | | 15  (100) | 15  (100) | 0  (0, 0-14) | 5  (33, 12-62) | 0  (0, 0-14) | 7  (47, 21-73) | 1  (7, 0-32) | 1  (7, 0-32) | 0  (0, 0-14) | 0  (0, 0-14) | 0  (0, 0-14) |
| Baseball  (n = 10) | | 10  (100) | 0  (0, 0-21) | 1  (10, 0-45) | 0  (0, 0-21) | 0  (0, 0-21) | 4  (40, 12-74) | 0  (0, 0-21) | 0  (0, 0-21) | 0  (0, 0-21) | 0  (0, 0-21) | 0  (0, 0-21) |

All data are reported as count (%, 95% confidence interval)

*Replication is not shown as no studies performed a replication

BJSM = British Journal of Sports Medicine; JSHS = Journal of Sport and Health Science; AJSM = American Journal of Sports Medicine; MSSE = Medicine Science Sport and Exercise

RCT = Randomized Controlled Trial
